# Medical Exemptions to Vaccination — Riverside County, California, 2016–2019

**DOI:** 10.1101/2022.04.14.22273891

**Authors:** Cameron Kaiser, Tonya Geiger, Cassandra Lynch, Anna Rivera

**Author notes:** Corresponding author: C. Kaiser, MD, MPH.

## Abstract

**Objectives:** To review the nature and clinical reasoning of medical exemptions to vaccination received in Riverside County, CA after California Senate Bill 277 eliminated personal belief exemptions statewide.

**Methods:** 614 deduplicated medical exemptions to vaccination from 156 providers were reviewed from August 2016 to August 2019. Exemptions covering all vaccines were additionally coded for number and category of medical justification.

**Results:** 81.3% of reviewed exemptions were for all vaccines, 91.0% were permanent or indefinite, and 74.9% were for all vaccines and permanent or indefinite. Of the 490 evaluated all-vaccine exemptions, a median of two and maximum of ten justifications were cited per exemption, most often a family history of autoimmune disease other than allergy. Three providers wrote more than 70 exemptions each.

**Conclusions:** The number and nature of the exemptions reviewed here raise concerns over their impact on immunization rates, and future policies may need to ensure additional oversight.

## Introduction

On July 1, 2016, California Senate Bill (SB) 277 eliminated personal belief exemptions to vaccination requirements for school and day care entry, and expanded the definition of medical vaccination exemptions to allow consideration of family history instead of only the student ‘s medical history (*1*). At the time of passage a medical exemption consisted of a licensed physician ‘s statement listing exempted vaccines and duration; neither specific reasoning nor renewal was required (*1*). Although exemptions overall decreased after SB 277 ‘s passage, medical exemptions increased statewide (*2,3,4,5,6,7*), including a modest increase in Riverside County (*4*), California ‘s fourth largest county with over 2.4 million residents (*8*). To evaluate the number and characteristics of medical exemptions after passage of SB 277, we conducted a study during August 2016–August 2019 of exemptions submitted to Riverside County educational institutions subject to the new requirements.

## Methods

Medical exemptions and grandfathered personal belief exemptions received for kindergarten and <u>7</u>^th^ grade entry and school transfers were sent from participating educational institutions by secure fax. Students ‘ names and identifying personal information were redacted in compliance with applicable state law and the federal Family Educational Rights and Privacy Act (FERPA). Duplicates were eliminated by comparing any school-assigned tracking numbers, letter date, provider name, justifications, and the document ‘s appearance, wording, or distinguishing marks.

Once records were deduplicated, the exemption date and the issuing provider ‘s name were recorded, along with a notation of whether the exemption was a grandfathered personal belief exemption or a current medical exemption. All exemptions submitted by schools during the study period were accepted for analysis, including those dated prior to it. Grandfathered personal belief exemptions were tallied and not further analyzed. Among medical exemptions, the following were documented: 1) whether the issuing provider was in the six-county (Riverside, San Bernardino, Los Angeles, Orange, San Diego and Imperial) region (defined as local), 2) whether the exemption was permanent or indefinite; and 3) whether the exemption covered all vaccines or a subset. These six counties were defined as local to avoid falsely labeling exemptions as out of the area for students receiving care at pediatric specialty hospitals in those jurisdictions; because students were not identified, their primary care providers therefore could not be known.

An exemption was considered permanent or indefinite if no expiry date was written, the provider explicitly declined to provide an end date, the letter explicitly indicated the exemption was permanent, or the provider indicated that the exemption was in force until the student ‘s 18 ^th^ birthday, as no school-required vaccinations would likely be administered after that time. An exemption was considered to cover potentially all vaccines if the provider explicitly wrote all vaccines were exempted, the provider used a checkbox form with required California immunizations and checked all listed vaccines, or the provider listed all subsequent required immunizations as being exempt.

Medical exemptions that appeared to cover all vaccines were reviewed by the county public health officer, a licensed family physician certified by the American Board of Family Medicine, who hand-coded justifications cited in the exemptions for analysis. Justifications were grouped where appropriate based on number and specificity of the condition(s).

At the study ‘s conclusion, all records of exemptions, intermediate work products, and associated documentation were securely destroyed. The study design and protocol were reviewed with the county health system Institutional Review Board (IRB), who determined IRB approval was not required.

## Results

Overall, 645 exemptions to vaccination were received during the study period; 31 were grandfathered personal belief exemptions, and 614 were medical exemptions. All 614 medical exemptions came from 156 providers, with 147 (94.2%) writing fewer than ten exemptions each and three (1.9%) writing more than 70 each (Table 1).

**TABLE 1.** Number of medical vaccine exemptions per provider, total number of medical exemptions written, and average number written by each provider — Riverside County, California, August 2016–August 2019.

| No. of medical<br>exemptions written<br>per provider | No. (%) of 614<br>total exemptions<br>written | No. (%) of 156<br>providers who wrote<br>exemptions | Average no.<br>exemptions written<br>per provider |
| --- | --- | --- | --- |
| <10 | 243 (39.6) | 147 (94.2) | 1.7 |
| 10–20 | 48 (7.8) | 3 (1.9) | 16.0 |
| 21–30 | 68 (11.1) | 3 (1.9) | 22.7 |
| 31–70 | 0 (0) | 0 (0) | 0 |
| 71–80 | 78 (12.7) | 1 (0.6) | 78.0 |
| >80 | 177 (29.2) | 2 (1.3) | 88.5 |

606 (98.7%) of the 614 medical exemptions were written by licensed physicians, with five by nurse practitioners, two by registered nurses, and one by a chiropractor. 478 (77.9%) of the medical exemptions were written by 103 providers (66.0%) outside of Riverside county, 453 (73.8%) were written by 65 providers (41.7%) outside of Riverside and San Bernardino counties, and 77 (12.5%) were written by 18 providers (11.5%) outside of all six counties. 131 (21.3%) of the medical exemptions were dated before the study start of August 2016. The highest individual school year count was 204 medical exemptions during the 2017-18 school year (Figure).

**FIGURE.**
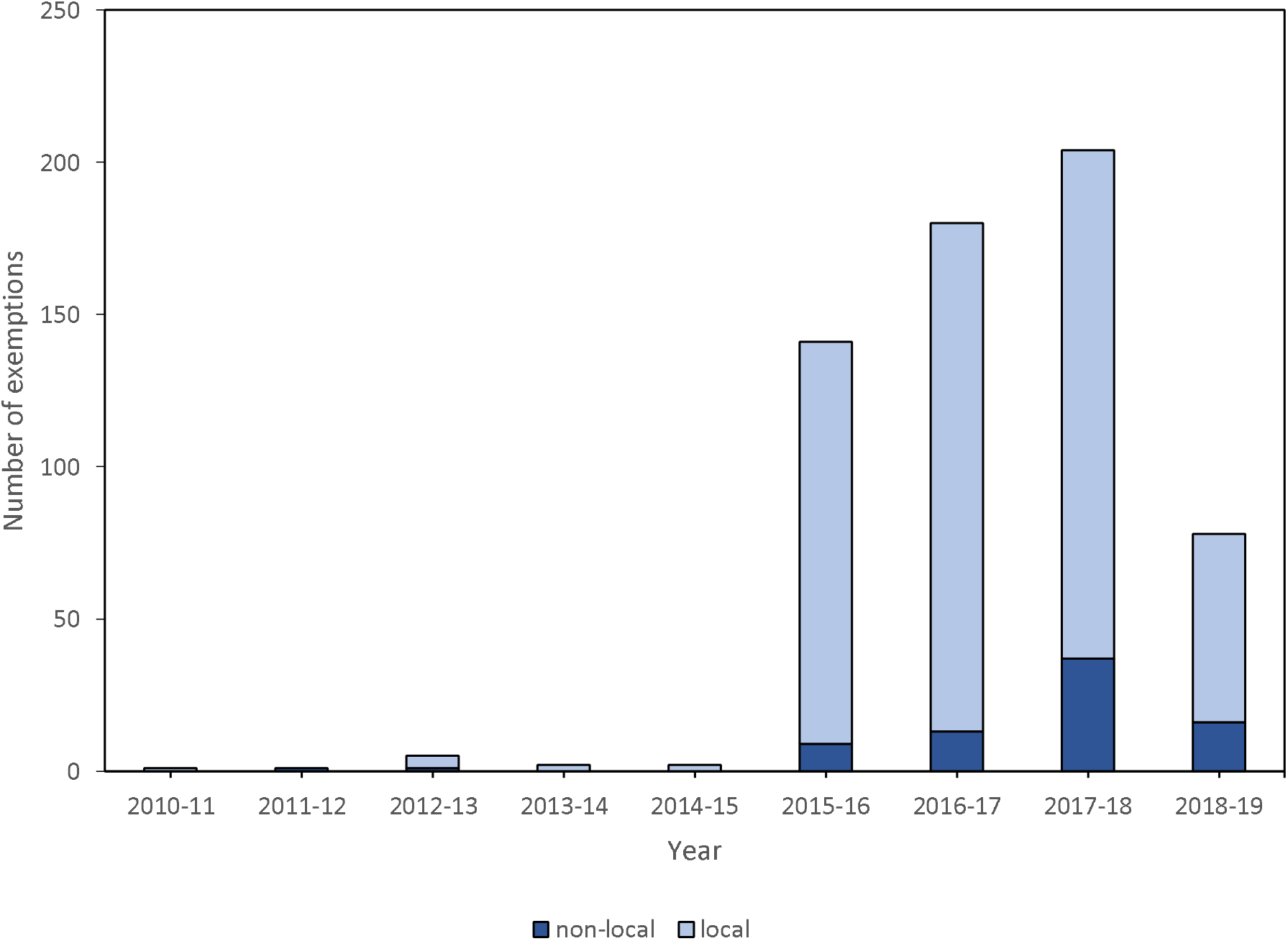
Issue dates of received medical exemptions by school year -- Riverside County, California (1) (2) (1) Senate Bill 277 effective July 1, 2016. (2) Senate Bills 276 and 714 passed September 9, 2019 and effective January 1, 2021.

Overall, 499 (81.3%) medical exemptions were for all vaccines, 559 (91.0%) were permanent or indefinite, and 460 (74.9%) were for all vaccines and were permanent or indefinite. Among the 499 exemptions for all vaccines, 490 exemptions (98.2%) were reviewed for justifications with nine discarded for illegibility. Nineteen (3.9%) of the 490 all-vaccine exemptions cited no justification for the exemption; all nineteen had no expiration date. The 471 exemptions containing justifications cited an average of 2.8 justifications per exemption, with a median of 2 and a maximum of 10 (Table 2). The most commonly cited justifications were family history of autoimmune disease other than allergy (263, 55.8%), family history of vaccine reaction (189, 40.1%), and history of vaccine reaction in the student (169, 35.9%). Autism spectrum disorder was cited in 37 (7.9%), with 22 asserting family history of autism spectrum disorder placing the student at risk, rather than history of autism spectrum disorder in the student. Overall, 152 (32.3%) of the 470 coded all-vaccine exemptions entirely cited family history and none of the student ‘s medical history.

**TABLE 2.** Distribution of coded justification and justification groups of medical exemptions to all vaccinations (n = 471) — Riverside County, California, 2016–2019 (1)

| **Justification or justification group** | **No. (%) of exemptions** |
| --- | --- |
| **Family history-based** |  |
| Family history of autoimmune disorders other than allergy | 263 (55.8) |
| Family history of vaccine reaction, sequelae or allergy | 189 (40.1) |
| Family history of neurologic/developmental disorder other than ASD (2) | 145 (30.8) |
| Family history of allergy other than vaccine allergy or reaction | 124 (26.3) |
| Family history of psychiatric disorder | 76 (16.1) |
| Unspecified statement of “family history” or “genetic susceptibility” | 24 (5.1) |
| Family history of ASD (2) | 22 (4.7) |
| Family history of malignancy | 17 (3.6) |
| Family history of gastrointestinal disorders other than autoimmune | 5 (1.1) |
| Family history of Lyme disease | 1 (0.2) |
| Family history of Guillain-Barre syndrome | 1 (0.2) |
| Family history of MTHFR (3) genetic abnormality | 1 (0.2) |
| **Student medical history-based** |  |
| History of vaccine reaction, sequelae or allergy | 169 (35.9) |
| History of neurologic or developmental disorder other than ASD (2) | 63 (13.4) |
| History of allergy other than vaccine allergy or reaction | 52 (11.0) |
| Unspecified general statement of “medical history” | 43 (9.1) |
| MTHFR (3) genetic abnormality | 42 (8.9) |
| History of autoimmune disorders other than allergy | 40 (8.5) |
| History of ASD (2) | 15 (3.2) |
| History of psychiatric disorder | 6 (1.3) |
| Drug-induced immunosuppression, including transplant recipients | 5 (1.1) |
| Testing in progress | 5 (1.1) |
| History of recurrent upper respiratory infections | 4 (0.8) |
| History of cardiac disease other than transplant recipient | 2 (0.4) |
| Already immune due to positive titer or other testing | 2 (0.4) |
| Chromosomal duplication 15q syndrome | 2 (0.4) |
| History of immunodeficiency | 2 (0.4) |
| Failure to thrive | 1 (0.2) |
| Acute disseminated encephalomyelitis | 1 (0.2) |
| History of recent infection not otherwise specified | 1 (0.2) |
| Gastroschisis | 1 (0.2) |
| History of malignancy | 1 (0.2) |

(1) Exemptions may contain multiple justifications.
(2) Autism spectrum disorder.
(3) Methylenetetrahydrofolate reductase.

## Discussion

This study shows a large increase in local medical exemption prevalence after passage of SB 277, consistent with state data showing a doubling of medical exemptions in the 2016-17 school year compared to 2015-16 (*2,3*), particularly in private schools, and continuing to increase through at least 2020 (*5,6,7*). Although legitimate contraindications exist for specific vaccinations (*9*), particular attention was paid to medical exemptions excluding all vaccines as there would rarely be a reason for all vaccines to be contraindicated (*9,10*) yet such exemptions were the majority of those received. Additionally, concern arises with the proportion of all-vaccine exemptions citing only family history, which has very few specific contraindications in the ACIP guidelines (*10*).

Because of anecdotal reports that some out-of-area providers publicly indicated willingness to write medical exemptions, non-local providers were also counted separately. The proportion of non-local providers who wrote medical exemptions may indicate at least some travel to providers outside the local area was undertaken to obtain them.

It is not known precisely what proportion of issued medical exemptions in Riverside County was actually collected by this study. Since identifying information was redacted, it is unknown if multiple exemptions covered the same student in multiple years, although this is not believed to be a significant factor as renewal was not required for permanent exemptions at the time of the study. School participation was voluntary, and some private schools did not submit any exemptions. Many exemptions contained directions forbidding disclosure to third parties; while neither California nor federal law prohibits health departments from obtaining deidentified information on vaccine exemptions, it is unclear how many were not sent to the health department because of such notations. It is also not known how many exemptions were subsequently rejected by schools for other reasons, although it is believed the vast majority remain in effect.

The study was concluded due to the imminent passage of California Senate Bills 276 and 714, both signed on September 9, 2019, which require periodic renewal of all medical exemptions, enable them to be revoked under certain conditions, and require them to be submitted to a central database. The bills also continue to permit medical exemptions outside of ACIP guidelines, including those based on family history, and allow revocations to be appealed (*11,12*).

Despite the bills ‘ passage, the California Department of Public Health reported in response to a media inquiry that as of October 2021 only 6% of medical exemptions were rejected statewide under these statutes (*13*). It is possible the new laws and other related enforcement actions have reduced the number of providers willing to write medical exemptions, including problematic ones; additionally, the numbers in this study may be peculiar to this region of the state, and the study does not claim to have exhaustively collected all medical exemptions issued in the county during the study period. However, the proportion of concerning exemptions in this set was apparently much greater than the amount rejected under statute thus far, and other policy reviews have highlighted the need for future monitoring of medical exemptions (*14*).

The number and nature of the medical exemptions reviewed here raise concerns over their impact on immunization rates, especially with respect to the current climate around COVID-19 immunization. When personal belief exemptions were eliminated, the number of medical exemptions increased dramatically in this study, making it an important determiner of community immunity. Additionally, the medical reasoning used for many of the exemptions reviewed here did not adhere to evidence-based standards. Future school vaccination policies may need to ensure regulatory oversight of medical exemptions so that students who legitimately cannot receive vaccines will remain protected by the students who do.

## Data Availability

All data produced in the present study are available upon reasonable request to the corresponding author except for all records of individual exemptions, intermediate work products, and associated documentation, which were securely destroyed.

